# Prenatal Prediction of Delayed Abdominal Closure in Isolated Congenital Diaphragmatic Hernia Using Observed-to-Expected Lung-to-Head Ratio

**DOI:** 10.64898/2026.11.04.26367256

**Authors:** Lucas Moratilla-Lapeña, José Luis Encinas, Eugenia Antolín, Ana Sánchez Torre, Francisco Hernández

## Abstract

**Objective:** To determine whether the observed-to-expected lung-to-head ratio (O/E LHR) independently predicts the need for delayed abdominal wall closure (DAC) in isolated left-sided congenital diaphragmatic hernia (CDH), and to establish an optimal cut-off value.

**Design:** Retrospective, single-centre cohort study.

**Setting:** Tertiary referral centre, Hospital Universitario La Paz, Madrid, Spain, January 2015 to May 2026.

**Patients:** 45 neonates with isolated left-sided CDH; 28 underwent primary abdominal closure and 17 required DAC.

**Interventions:** None (observational study).

**Main outcome measures:** Need for DAC; discriminative capacity of O/E LHR for DAC and its optimal cut-off; O/E LHR–DAC association adjusted for liver herniation.

**Results:** DAC was associated with lower gestational age at birth (37.00 [34.00–38.00] vs 39.00 [38.00–39.00] weeks), lower birth weight (2,690 [2,290–3,210] vs 3,120 [2,968– 3,285] g), higher rates of liver herniation (13/17 vs 1/28), greater ECMO requirement (9/17 [52.9%] vs 3/28 [10.7%]), and higher in-hospital mortality (35.3% vs 3.6%) than primary closure. O/E LHR showed excellent discriminative ability for DAC (AUC 0.939, 95% CI 0.877 to 1.000), with an optimal cut-off of 34.1% (sensitivity 82.4%, specificity 82.1%, positive predictive value 73.7%, negative predictive value 88.5%). O/E LHR remained independently associated with DAC after adjustment for liver herniation (adjusted OR 0.82 per one-point increase, 95% CI 0.67 to 0.94).

**Conclusions:** O/E LHR was independently associated with the risk of DAC beyond the effect of liver herniation. These preliminary findings suggest a role for O/E LHR in prenatal risk stratification for DAC, pending external validation in larger, multicentre cohorts.

**What’s already known about this topic?:** O/E LHR is an established prenatal predictor of survival and pulmonary morbidity in isolated CDH. Delayed abdominal closure has been linked to lower LHR, liver herniation, and higher ECMO use and mortality, but without adjusting for confounders or using O/E LHR.

**What does this study add?:** O/E LHR independently predicts DAC risk beyond liver herniation, with excellent discriminative ability (AUC 0.939) and a proposed cut-off of 34.1%, extending its prognostic role to this surgical outcome.

**Data availability statement:** The data that support the findings of this study are available from the corresponding author upon reasonable request.

**Note:** This study is based on a cohort that partially overlaps with our previously published report on delayed dynamic abdominal wall closure using a Gore-Tex patch ^1^ , which addressed surgical technique and outcomes. The present study addresses the distinct question of prenatal predictors of the need for delayed closure. This overlap is disclosed in accordance with ICMJE recommendations.

## 1. Introduction

Congenital diaphragmatic hernia (CDH) is a major structural anomaly with a prevalence around 2.3 per 10000 live births. Despite substantial advances in perinatal and neonatal management, survival remains limited to 60–70% even in high-volume referral centers, reflecting pulmonary hypoplasia and hypertension ^2^.

The observed-to-expected lung-to-head ratio (O/E LHR), first described by Jani et al.^3^, expresses the fetal LHR as a percentage of the value expected for gestational age, overcoming the inherent gestational-age dependence of the raw LHR and allowing comparison across pregnancies assessed at different points in gestation. O/E LHR has been consistently validated as a predictor of neonatal survival in isolated CDH, although its prognostic value has been established almost exclusively in relation to survival and early pulmonary morbidity ^4,5^, while its relationship with specific postoperative surgical outcomes remains far less characterized.

Beyond the diaphragmatic defect itself, the ability to achieve fascial closure of the abdominal wall carries prognostic significance of its own: when primary fascial approximation is not feasible and an abdominal wall patch is required instead, mortality has been reported to be substantially higher than after primary fascial closure ^6^. Yet the handful of studies that have specifically examined delayed abdominal closure (DAC) offer a discordant picture of its prenatal predictors. Laje et al.^7^ found that patients requiring DAC had lower LHR values, a higher rate of liver herniation, and higher mortality than those undergoing primary closure. In contrast, Maxwell et al.^8^, analyzing the CAPSNet registry, found a higher rate of liver herniation among patients requiring DAC, but no difference in LHR nor in mortality, despite a clear association with postoperative morbidity.

Notably, neither study reported the O/E LHR: both relied on the raw LHR a less standardized severity marker than O/E LHR. We previously described a single-center cohort in which patients requiring DAC had markedly lower O/E LHR and a higher rate of liver herniation than those undergoing primary fascial closure ^1^, but this observation was purely descriptive and unadjusted for potential confounders. Taken together, whether O/E LHR independently predicts the need for DAC remains unknown. We therefore aimed to determine the predictive capacity of O/E LHR for the need of DAC in a cohort of isolated left-sided CDH and to establish an optimal cut-off value to assist in preoperative surgical planning.

## 2. Methods

### 2.1. Study design and population

We conducted a retrospective, single-center study of neonates with CDH treated at Hospital Universitario La Paz, Madrid, Spain, between January 2015 and May 2026. Only neonates with isolated CDH, defined as the absence of major associated structural anomalies or a confirmed chromosomal abnormality, were included.

Inclusion criteria were: (1) prenatal assessment of CDH at our center; (2) surgical repair of CDH during the neonatal period (age ≤ 28 days); and (3) admission to the Neonatal Intensive Care Unit (NICU).

Exclusion criteria were: (1) absence of prenatal diagnosis; (2) absence of an O/E LHR measurement within the 25+0 − 29+6 weeks’ gestation window; (3) presence of right-side CDH given the distinct normal reference ranges and clinical course described for right-sided defects^9^ and (4) death prior to surgical correction.

The study was conducted in accordance with the Declaration of Helsinki and approved by the Institutional Ethics Committee of Hospital Universitario La Paz (PI-6678 2025.317, 14 May 2025).

### 2.2. Data collection and study variables

Data were collected through retrospective review of electronic medical records. The following variables were recorded: demographic data (sex, gestational age at birth, birth weight); prenatal severity markers (O/E LHR, O/E LHR severity category, liver position); fetal therapy (use of fetoscopic endoluminal tracheal occlusion [FETO]); neonatal condition at birth (Apgar score at 1 and 5 minutes); respiratory and hemodynamic support (need for high-frequency oscillatory ventilation [HFOV]; need for extracorporeal membrane oxygenation [ECMO]); surgical variables (need for DAC); and postoperative outcome (in-hospital mortality). Patients were grouped according to need for DAC or primary abdominal closure during CDH repair.

### 2.3. Prenatal assessment

Fetal lung volume was assessed by ultrasound measurement of the lung-to-head ratio (LHR) at 25-29 weeks of gestation in line with the original description of prenatal severity assessment in CDH ^10^, performed by the two same obstetricians. The observed LHR was expressed as a percentage of the expected LHR for gestational age (O/E LHR), calculated using the reference formula and normative values described by Jani et al ^3^. Liver position was recorded for each case as a well-established prognostic factor ^11^.

O/E LHR severity was categorized according to Deprest et al. ^5^ as extreme (<15%), severe (15–25%), moderate (26–35%) or mild (≥36%) in fetuses without liver herniation, and as extreme (<15%), severe (15–25%), moderate (26–45%) or mild (≥46%) in fetuses with liver herniation. In the subset of fetuses who underwent FETO, the baseline O/E LHR value obtained prior to balloon insertion was used for analysis, to reflect the native severity of pulmonary hypoplasia rather than the treatment-induced lung growth response.

### 2.4. Postnatal management

Diaphragmatic hernia repair was performed via an open surgical approach through a left subcostal incision or through a thoracoscopic approach. We defined DAC as any case in which the abdominal wall was not closed at the time of diaphragmatic hernia repair, requiring a separate, subsequent closure procedure; conversely, patients underwent primary fascial closure when the abdominal wall was closed during the same operation. The decision to defer abdominal wall closure was guided by intraoperative findings, consistent with the institutional criteria previously reported by our group ^1^: closure was delayed either when fascial approximation could not be achieved, or when intra-abdominal pressure after attempted closure, measured via a bladder catheter, exceeded 10 mmHg, the pediatric threshold for intra-abdominal hypertension defined by the World Society of Abdominal Compartment Syndrome consensus guidelines ^12^ .

### 2.5. Statistical analysis

Statistical analysis was performed using RStudio 2024.12.0 (Posit Software, PBC) within the R environment (version 4.5.3). The normality of data distribution was assessed using the Shapiro-Wilk test. Continuous variables are presented as median and interquartile range (IQR), while categorical variables are expressed as absolute frequencies and percentages. Categorical variables were compared between groups using Fisher’s exact test, whereas continuous variables were compared using the Wilcoxon rank-sum test. The discriminative capacity of O/E LHR for predicting DAC was assessed by receiver operating characteristic (ROC) curve analysis, with the area under the curve (AUC) and its 95% confidence interval estimated using *pROC package*. Internal validation of the discriminative performance of O/E LHR was performed using Harrell’s bootstrap optimism-correction method (1000 resamples; *rms package*).

The optimal O/E LHR cut-off was determined using a bootstrap-aggregated approach (*cutpointr package*), in which the cut-off minimizing a joint sensitivity/specificity criterion was re-estimated across 1000 bootstrap resamples and aggregated to obtain a robust threshold. Sensitivity, specificity, positive predictive value and negative predictive value were reported both as apparent values (calculated in the full sample at the aggregated cut-off) and as optimism-corrected values, obtained from the out-of-bag performance across the same 1000 resamples and summarized as the mean and 95% percentile interval across replicates.

The discriminative performance of the continuous O/E LHR was compared with O/E LHR severity category for the prediction of DAC, using a paired DeLong test to compare the two areas under the curve. Logistic regression was used to estimate univariate odds ratios (OR) for O/E LHR as a predictor of DAC, followed by a multivariate model adjusting for liver position (liver-up). Given the small sample size, multivariate logistic regression was performed using Firth’s penalized maximum-likelihood method, which provides less biased effect estimates and more stable confidence intervals than standard maximum-likelihood logistic regression when sparse-data or separation issues are present. The potential confounding or mediating role of ECMO was assessed using Fisher’s exact and Wilcoxon rank-sum tests, followed by a post-hoc sensitivity analysis restricting the ROC and AUC analysis to patients who did not require ECMO. A two-sided p-value <0.05 was considered statistically significant.

## 3. Results

During the study period, 67 patients with CDH were initially assessed, of whom 45 met the inclusion criteria and were included in the final analysis. Twenty-two patients were excluded: four died before surgical repair (mean O/E LHR 22.1%, range 19–26%; one while on ECMO and three who were not ECMO candidates), seven had a postnatal diagnosis of CDH without a prenatal diagnosis, six had late prenatal diagnosis (≥30 weeks) and five were right sided CDH.

The final cohort had a total of 17 (37.8%) patients who underwent DAC and 28 (62.2%) patients whose abdomen was closed primarily after CDH repair. Baseline demographic and prenatal characteristics of the cohort, stratified by the need for DAC, are summarized in Table 1. There were no significant differences between the DAC and primary closure groups in sex distribution. Gestational age at birth and birth weight were lower among patients requiring DAC than those who underwent abdominal primary closure (*p < 0.001* and *p = 0.017*).

**Table 1.**
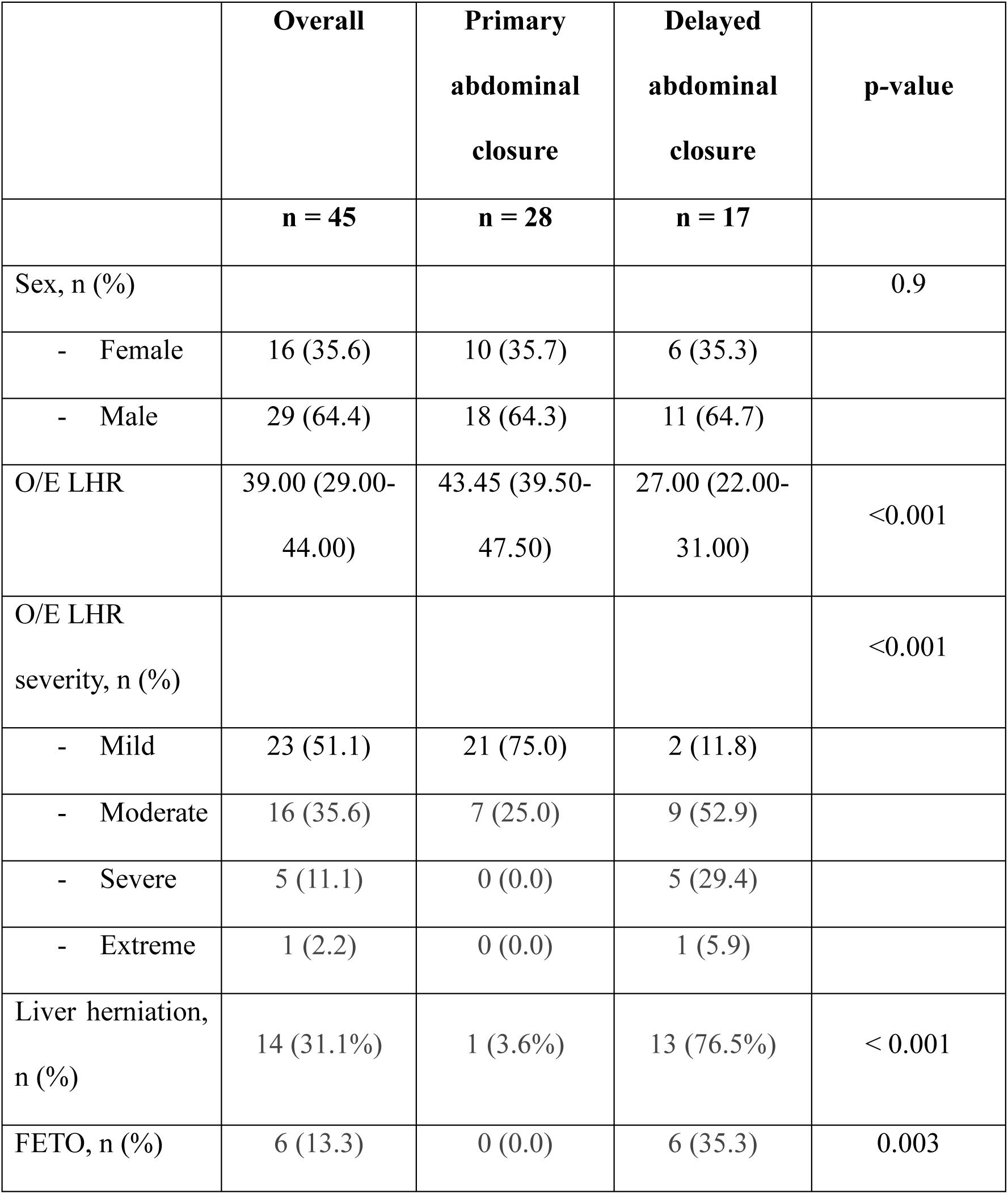
Prenatal baseline characteristics according to need for delayed abdominal closure.

|  | **Overall** | **Primary abdominal closure** | **Delayed abdominal closure** | **p-value** |
|---|---|---|---|---|
|  | **n = 45** | **n = 28** | **n = 17** |  |
| Sex, n (%) |  |  |  | 0.9 |
| - Female | 16 (35.6) | 10 (35.7) | 6 (35.3) |  |
| - Male | 29 (64.4) | 18 (64.3) | 11 (64.7) |  |
| O/E LHR | 39.00 (29.00-44.00) | 43.45 (39.50-47.50) | 27.00 (22.00-31.00) | <0.001 |
| O/E LHR severity, n (%) |  |  |  | <0.001 |
| - Mild | 23 (51.1) | 21 (75.0) | 2 (11.8) |  |
| - Moderate | 16 (35.6) | 7 (25.0) | 9 (52.9) |  |
| - Severe | 5 (11.1) | 0 (0.0) | 5 (29.4) |  |
| - Extreme | 1 (2.2) | 0 (0.0) | 1 (5.9) |  |
| Liver herniation, n (%) | 14 (31.1%) | 1 (3.6%) | 13 (76.5%) | < 0.001 |
| FETO, n (%) | 6 (13.3) | 0 (0.0) | 6 (35.3) | 0.003 |

According to prenatal assessment, patients requiring DAC had a significantly lower O/E LHR than those undergoing primary closure (27.00 [22.00-31.00] vs 43.45 [39.50-47.50]; *p < 0.001*). Accordingly, the distribution of O/E LHR severity categories differed significantly between groups (*p < 0.001*), with a higher proportion of extreme and severe cases among patients requiring DAC (Table 1). Intrathoracic liver herniation was also more frequent in the DAC group (13/17 vs 1/28, *p < 0.001*). FETO was performed in 6 patients, all of whom belonged to the DAC group.

Neonatal and perioperative characteristics, stratified by the need for DAC, are summarized in Table 2. Apgar scores at 1 and 5 minutes did not differ significantly between groups (*p = 0.6* and *p = 0.076*). The need for HFOV (*p = 0.006*) and ECMO (*p = 0.004*) was significantly more frequent among patients requiring DAC. In-hospital mortality was also significantly higher in the DAC group (35.3% vs. 3.6%, *p = 0.008*). All 45 patients underwent open surgical repair of the diaphragmatic defect; no cases were approached thoracoscopically.

**Table 2.** Perinatal baseline characteristics according to need for delayed abdominal closure.

|  | **Overall** | **Primary abdominal closure** | **Delayed abdominal closure** | **p-value** |
|---|---|---|---|---|
|  | **n = 45** | **n = 28** | **n = 17** |  |
| Gestational age at delivery (weeks) | 38.00 (37.00-39.00) | 39.00 (38.00-39.00) | 37.00 (34.00-38.00) | <0.001 |
| Birth weight (g) | 3,030 (2,690-3,240) | 3,120 (2,968-3,285) | 2,690 (2,290-3,210) | 0.017 |
| APGAR 1 | 5.00 (3.00-7.00) | 5.00 (3.00-8.00) | 4.00 (3.00-7.00) | 0.6 |
| APGAR 5 | 8.00 (6.00-9.00) | 8.00 (6.00-9.00) | 7.00 (5.00-8.00) | 0.076 |
| HFOV, n (%) | 22 (48.9) | 9 (32.1) | 13 (76.5) | 0.006 |
| ECMO, n (%) | 12 (26.7) | 3 (10.7) | 9 (52.9) | 0.004 |
| Mortality, n (%) | 7 (15.6) | 1 (3.6) | 6 (35.3) | 0.008 |

### 3.1. Discriminative capacity of O/E LHR and optimal cut-off for DAC

The O/E LHR showed good discriminative capacity for predicting the need for DAC, with an AUC of 0.939 (95% CI, 0.877-1.000). Internal validation using Harrell’s bootstrap method (1000 resamples) showed minimal optimism, with a corrected AUC of 0.938 and satisfactory calibration (corrected slope 0.926, corrected Brier score 0.119) (Supplementary Figure 1).

The optimal O/E LHR cut-off value, determined using a bootstrap-aggregated approach across the same 1000 resamples, was 34.1 (Fig. 1). Apparent and optimism-corrected performance metrics were closely consistent (Table 3), indicating minimal overfitting despite the limited sample size.

**Figure 1.**
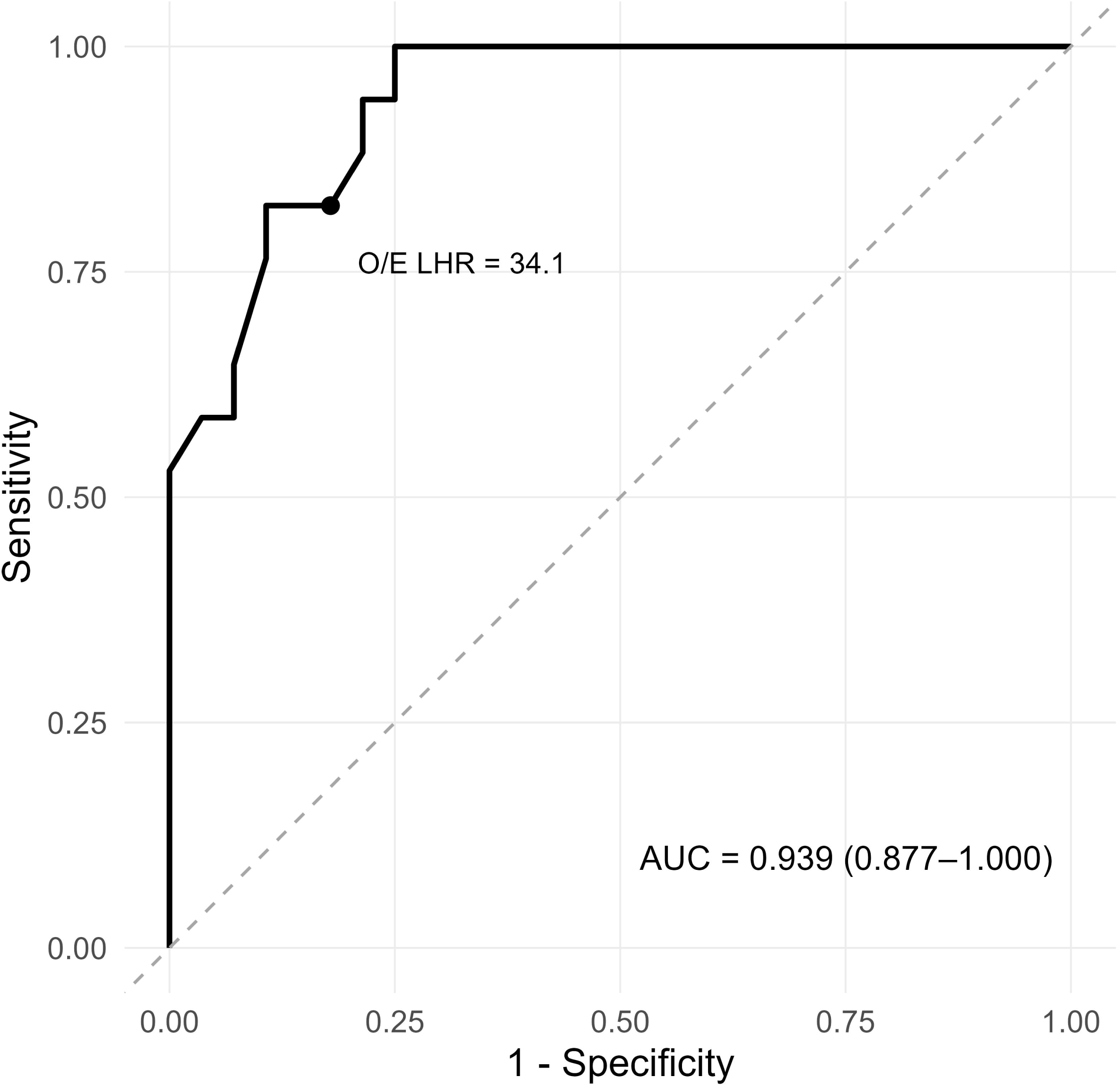
ROC curve for the continuous O/E LHR as a predictor of delayed abdominal wall closure DAC. AUC, 0.939 (95% CI, 0.877–1.000). The dashed line represents the line of no discrimination. The dot marks the optimal cut-off (O/E LHR = 34.1%; sensitivity, 82.4%; specificity, 82.1%).

**Table 3.** Performance of the O/E LHR cut-off (34.1%) for DAC.

| **Metric** | **Apparent** | **Optimism-corrected (95% CI)** |
|---|---|---|
| Sensitivity | 82.35% | 81.2 (33.3-100) % |
| Specificity | 82.14% | 83.6 (57.1-100) % |
| Positive predictive value | 73.68% | 75.9 (42.9-100) % |
| Negative predictive value | 88.46 % | 89.0 (66.7-100) % |
| AUC | 0.939 | 0.942 (0.833-1.00) |

When comparing the continuous O/E LHR with its severity category classification, the former showed significantly higher discriminative ability for DAC than the latter (AUC, 0.860; 95% CI, 0.755–0.966), with a difference in AUC of 0.079 (95% CI, 0.004–0.154; *p = 0.039*) (Fig. 2). This finding supports the use of the continuous O/E LHR, rather than the pre-existing severity classification, for the prediction of DAC.

**Figure 2.**
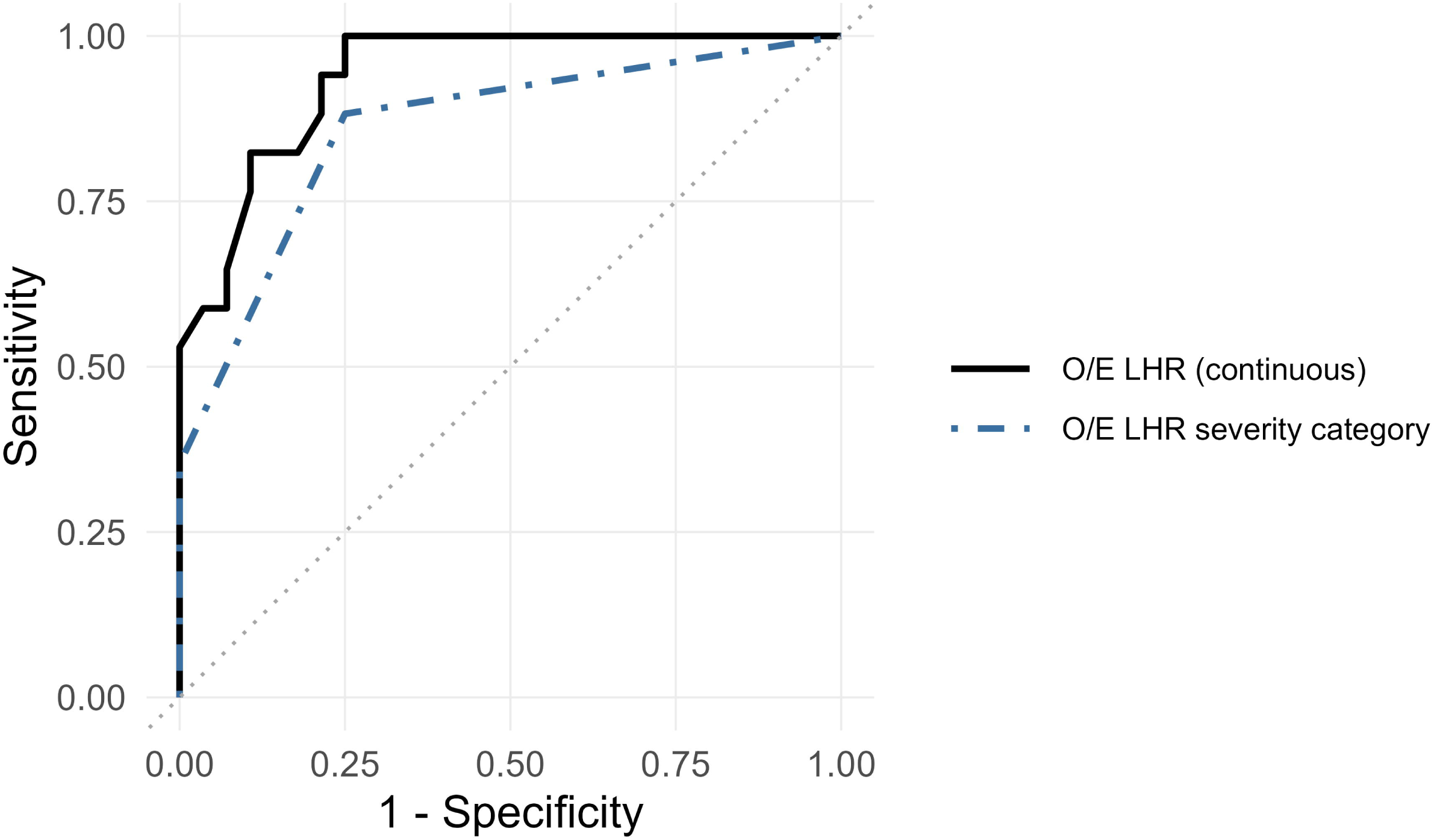
Comparison of ROC curves for the continuous O/E LHR and its severity category classification as predictors of DAC. Continuous O/E LHR (solid line; AUC, 0.939) versus severity category (dashed line; AUC, 0.860). The AUCs differed significantly (*p = 0.039*).

In a post-hoc sensitivity analysis restricted to the 33 patients who did not require ECMO (8 DAC events), O/E LHR discrimination for DAC remained high (AUC 0.923 [95% CI, 0.834-1.00], comparable to the overall cohort, though this subgroup estimate is imprecise given the small event count.

### 3.2. Association between O/E LHR and DAC: univariate and multivariate analysis

In univariate analysis, higher O/E LHR was significantly associated with lower odds of DAC (*p < 0.001*), corresponding to a 24.7% reduction in the odds of DAC per one-point increase. After adjustment for liver herniation using Firth’s penalized-likelihood logistic regression, the O/E LHR remained an independent predictor of DAC (*p = 0.002*), indicating a 17.9% reduction in the odds of DAC per one-point increase, while liver herniation was independently associated with higher odds of DAC (*p = 0.008*) (Supplementary Table 1).

## 4. Discussion

This study assessed the value of the continuous O/E LHR, compared with its conventional severity-category classification, for predicting the need for DAC in fetuses with isolated CDH. A key finding is the strong discriminative ability of the O/E LHR for DAC (AUC, 0.939), which outperformed the severity-category classification and remained associated with DAC after adjustment for liver herniation. DAC was, in turn, associated with a higher requirement for ECMO and HFOV support and with higher in-hospital mortality. Previous evidence has established the O/E LHR as an excellent prenatal measure to predict pulmonary hypoplasia and mortality ^2,3,5,10,13^ and severity grading based on O/E LHR and liver herniation has been used mainly to stratify mortality risk and select FETO candidates ^5,14,15^. MRI-based lung volume measures (TFLV and PPLV) have similarly predicted survival, pulmonary hypertension, or ECMO need ^16–19^, with recent work refining O/E LHR and O/E TFLV cut-offs for these outcomes ^20^. Fetal-MRI lung volume and liver herniation have also predicted the need for patch repair ^21,22^. However, to our knowledge none of these markers had previously been examined in relation to staged abdominal wall closure.

Addressing this gap, we found the continuous O/E LHR to be a strong predictor of DAC, with an optimal cut-off of 34.1% in our cohort, extending its established prognostic value beyond survival and respiratory morbidity to this specific surgical endpoint. This discriminative performance was confirmed by bootstrap internal validation and held-up in a post-hoc sensitivity analysis excluding ECMO cases, suggesting it was not substantially driver by overfitting or by ECMO related case severity alone, though precision in the ECMO excluded subgroup was limited by the small number of events.

When liver herniation was added as a covariate in the multivariable model, the O/E LHR remained associated with DAC (adjusted OR, 0.82 [95% CI, 0.67–0.94] per one-point increase, corresponding to a 17.9% reduction in the odds of DAC [95% CI, 6.1–33.2% reduction]). Liver herniation itself showed a strong association (OR, 14.57 [95% CI, 1.99–185.66]), consistent with its established prognostic role in CDH ^11,23^; however, the wide confidence interval around this estimate reflects the limited number of events in this subgroup and this specific estimate should be interpreted with caution. Even allowing for that imprecision, the combination of these two covariates carries a practical implication for severity classification. Under the commonly used risk stratification categories in CDH ^5^, our cohort’s DAC cut-off of 34.1% falls within the moderate range regardless of liver status, yet two fetuses classified identically as moderate risk may carry different odds of DAC depending on whether the liver is herniated or not. This may partly explain why the continuous O/E LHR outperformed the severity category classification in our ROC analysis, and support reporting O/E LHR as a continuous variable, with liver herniation as a separate covariate, rather than collapsing both into a single ordinal category, at least for DAC.

The clinical significance of DAC itself remains debated. Laje et al.^7^ found DAC far more common among CDH patients requiring ECMO, but within the ECMO subgroup, DAC versus primary closure did not differ in prenatal severity or survival, suggesting ECMO status, not closure strategy, drive the outcome. Maxwell et al.^8^ similarly linked DAC to right-sided defects and greater perioperative morbidity, without increased mortality. Together, these findings frame DAC as a marker of disease severity rather than an independent determinant of survival, consistent with our own prior report ^1^, in which DAC was associated with lower O/E LHR, more advanced defect type and greater ECMO requirement, with mortality reflecting this high-risk profile rather than the closure strategy itself.

Taken together, these findings suggest that O/E LHŔs established role in predicting mortality, ECMO need, and postoperative respiratory and feeding morbidity may extend to DAC risk as well. Recognizing this risk before birth may therefore help clinicians identify this distinct pattern of disease complexity early, informing parental counseling, the perioperative planning of staged abdominal closure, the anticipated need for prolonged nutritional and ventilatory support driven by the underlying disease severity, and the referral to an appropriately resourced center, informed by the likelihood of ECMO requirement that O/E LHR already predicts.

Despite these potential clinical implications, several considerations warrant caution before incorporating O/E LHR based prediction of DAC risk into routine perinatal counseling or management protocols. The model presented here was derived from a small, single-center cohort, and some of its estimates should be interpreted as hypothesis-generating rather than as an externally validated clinical tool. Furthermore, the specific O/E LHR cutoff identified for DAC risk in this cohort (34.1%) reflects the characteristics of this particular population, and its generalizability may be limited by variation across centers in surgical thresholds for staged closure, case-mix, and how DAC itself is operationally defined. External validation in larger, multicenter cohorts will therefore be needed before these findings can meaningfully inform individualized counseling or resource-allocation decisions.

This study has several limitations. It is a retrospective, single-center analysis of a relatively small cohort, which limits statistical power and despite the use of Firth’s penalized regression to address the quasi-complete separation observed in the multivariable model, still resulted in a wide confidence interval for the liver herniation estimate, reflecting the small number of events in this subgroup rather than a precise effect size. Additionally, because the four patients who died before surgical repair could not be assigned to a closure group, our model estimates DAC risk conditional on survival to the closure decision, not as an unconditional prenatal prediction. Similarly, confidence limits around the optimism-corrected estimates remained wide, reflecting the small number of DAC events and underscoring the preliminary nature of these estimates. This cohort partially overlaps with that of our previously published report on delayed dynamic abdominal wall closure^1^, an overlap disclosed here in accordance with ICMJE recommendations. Although the two studies address distinct questions, some shared patients were unavoidable. Finally, this cohort included only isolated left-sided CDH, limiting the generalizability of our findings to right-sided defects and to cases with associated anomalies.

## 5. Conclusion

In this retrospective, single-center cohort, O/E LHR was independently associated with the risk of DAC in isolated left-sided CDH, beyond the well-established effect of liver herniation. These preliminary findings suggest that O/E LHR may have a role in prenatal risk stratification for DAC, potentially complementing existing counseling and perioperative planning frameworks. However, external validation in larger, multicenter cohorts is needed before these findings can guide clinical decision-making.

## Data Availability

All data produced in the present study are available upon reasonable request to the authors

## Author Contribution Statement

Conceptualization, ML.L and E.JL..; Methodology, ML.L. and E.JL.; Software, ML.L; Validation, ML.L., E.JL., A.E., ST.A. and H.F.; Formal Analysis, ML.L.; Investigation, ML.L. and ST.A.; Resources, ML.L., A.E., ST.A. and E.JL.; Data Curation, ML.L.; Writing – Original Draft Preparation, ML.L.; Writing – Review & Editing, E.JL., A.E., ST.A. and H.F.; Visualization, E.JL., A.E., ST.A. and H.F.; Supervision, H.F.

## Patient consent statement

Patient consent statement is not applicable or needed for this study due to its retrospective nature,

## Declaration of generative AI and AI-assisted technologies in the manuscript preparation process

During the preparation of this work, the author(s) used Claude (Anthropic) in order to check and correct the grammar and language of the manuscript. After using this tool/service, the author(s) reviewed and edited the content as needed and take(s) full responsibility for the content of the published article.

