## Supplementary Figure 1; Supplementary Table 1 for "Prenatal Prediction of Delayed Abdominal Closure in Isolated Congenital Diaphragmatic Hernia Using Observed-to-Expected Lung-to-Head Ratio"

**Supplementary Figure S1. Calibration plot for the logistic regression model relating O/E LHR to DAC, based on 1000 bootstrap resamples (Harrell's method).** The dotted line shows apparent calibration; the solid line shows bias-corrected (optimism-corrected) calibration; the dashed line represents perfect calibration; grey lines denote 95% confidence limits. B = 1000 repetitions; n = 45; mean absolute error = 0.03.

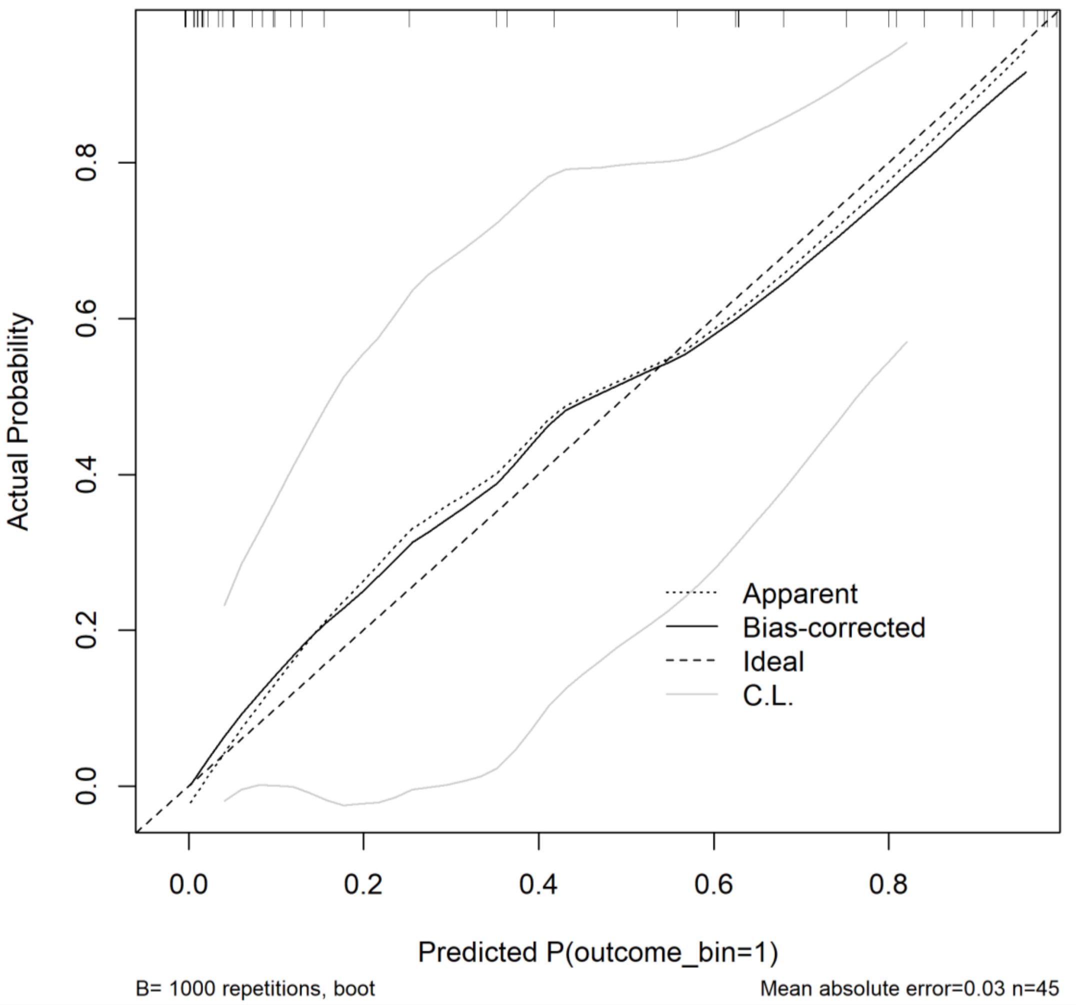

**Supplementary Table 1.** Logistic regression for DAC

| **Variable** | **OR** | **95% CI** | **p-value** |
|---|---|---|---|
| O/E LHR, univariate | 0.753 | 0.613-0.860 | <0.001 |
| O/E LHR, multivariate | 0.821 | 0.668-0.939 | 0.002 |
| Liver herniation, multivariate | 14.57 | 1.99-185.66 | 0.008 |
